# Willingness and Perceived ability to pay for Uganda’s Proposed National Health Insurance Scheme among Informal Sector workers in Iganga and Mayuge districts, Uganda: A Contingent Valuation Method

**DOI:** 10.1101/2024.07.24.24310952

**Authors:** Noel Namuhani, Angela Kisakye, Suzanne N Kiwanuka

## Abstract

**Background:** Access to health care remains a challenge, especially among the informal sector workers in most low-income countries, due to high out-of-pocket (OOP) expenditures, with Uganda spending over 28.0% out of pocket on health care. In response, Uganda has proposed a national health insurance scheme (NHI). However, the willingness and ability to pay for the proposed NHI scheme within the informal sector have not yet been explored in Uganda. This study assessed the willingness and perceived ability to pay for the proposed NHI scheme and its determinants among the informal sector workers in Iganga and Mayuge districts, Uganda.

**Methodology:** A cross-sectional study was conducted in Iganga and Mayuge districts in April and May 2019. A contingent valuation method using the bidding game technique was used to elicit the willingness to pay (WTP). A total of 853 informal sector workers, including farmers, commercial motorists, fishermen, and traders, were randomly sampled. Seven focus group discussions (FGD) were also conducted. Logistic regression was done to identify the determinants of willingness to pay for the proposed NHI scheme. Qualitative data was analyzed thematically.

**Results:** The majority 695/853, (81.5%) of the respondents were willing to pay for NHI; the median WTP was UGX 25,000 (USD 6.8) annually; and 633/853, (74.2%) of the respondents believed that they were able to pay for the health insurance. Willingness to Pay was significantly associated with being a fisher folk (AOR: 1.70 95%CI: 1.04-2.79, P = 0.035), being in the fourth wealth quintile (AOR: 2.98, 95% CI: 1.56–5.65), not hearing about health insurance (AOR: 0.50 95%CI: 0.23-0.86, P = 0.032), and not having saving group membership (AOR: 0.51, 95%CI: 0.34-0.76, P<0.001). Most of the FGD participants were willing to pay for the proposed scheme; however, some of the participants doubted their ability to pay for the scheme given their high poverty levels and their unstable income.

**Conclusion:** The willingness to pay for health insurance in the informal sector is high. Therefore, it is viable for the government to extend NHI to the informal sector. However, awareness building and due consideration of high poverty levels in setting appropriate premiums should be a priority.

## Introduction

Access to health care remains a big challenge, especially among the informal sector in most low-income countries (1, 2), mainly due to high out-of-pocket (OOP) expenditures (3). This put about 150 million people at risk of catastrophic healthcare costs annually, driving over 100 million people into poverty worldwide (3). According to the World Bank, direct modes of payment for health care services constitute 40% of health care expenditure in developing countries (4).

In Uganda, although the national health accounts reported a reduction in the out-of-pocket expenditure on health as a share of current health expenditure (CHE) from 38.6% in the financial year 2018/2019 to 27.4% in the financial year 2020/2021, the OOPE remains significantly high (5). Accessing healthcare is associated with several expenses, including transport, drugs, diagnostics, and consultancy fees, among other expenses related to personnel and capital costs (6).

In Uganda, over 80% of the labor force belongs to the informal sector (7). These include peasant farmers, commercial cyclists, fishermen, and those employed in unregistered or small-scale enterprises. It is estimated that 59% of the people in the informal sector in Eastern Uganda cannot afford to access quality health care due to high poverty levels and OOP for health care (8).

Globally, the implementation of health insurance schemes is one of the major responses to addressing out-of-pocket payments for health care (3, 9) in low and middle-income countries. Health insurance has the ability to reduce financial barriers to health care access and protect individuals and families against the risk of unpredictable health care expenditures, hence aiding the attainment of UHC (10). This has improved access to healthcare, especially in developed countries.

High coverage and enrollment in the health insurance scheme are critical for the success and sustainability of the scheme. However, enrolment in health insurance schemes remains low, ranging from 2% to 40% in most African countries (11–13). The poor enrolment rates in the different countries have been attributed to a number of factors, including low incomes, large household sizes, long distances to health facilities, limited awareness, poor quality of health care services, inappropriate benefit packages, a lack of trust in the systems, and high illiteracy levels, especially in the informal sector (14–16). The informal sector workers are sometimes of low socioeconomic status compared to the formal sector workers, hence having a lower ability to pay for health insurance (16). Furthermore, the informal sector is not well organized; they have unpredictable incomes, making it difficult to enroll and register them into the NHI scheme and collect regular contributions from them (13, 17).

Uganda is in the process of developing a NHI scheme to address the high OOP and access to services. However, given the high poverty levels among Ugandans and more so in the informal sector, the policy question of how much money people are willing to spend and their ability to pay remains unanswered. Eastern Uganda is the poorest region in Uganda, with 35.7% of most households living below the poverty line (7). The willingness to pay for the proposed NHI scheme, especially within the informal sector, which makes up 80% of Uganda’s population, is critical to the achievement of equity in access to care and the core risk pooling function of a health insurance scheme. This would translate into high enrollment rates, which foster the financial sustainability of the schemes (10).

The national health insurance bill, 2019, proposes that the scheme will be mandatory for all Ugandans, and each person is to pay a premium that will be determined by the board. The bill was passed by parliament in 2021 but was not assented to by the president due to disagreements among stakeholders who were not consulted. As of 2024, the bill is being discussed by the Ministry of Health and stakeholders before being re-tabled in the Parliament of Uganda.

There is a paucity of information on the willingness and ability to pay for the proposed NHI, especially within the informal sector. Therefore, this study aimed at documenting the willingness to pay and the perceived ability to pay for the proposed NHI and factors that influence willingness to pay for the proposed NHI scheme among the informal sector workers. This study’s findings will be used by the MOH in determining affordable premiums and how they should be paid.

## Materials and Methods

### Study design

This was a cross-sectional study design that involved mixed methods of data collection. This study adopted a contingent valuation method (CVM) using the bidding game technique to directly elicit or measure willingness to pay for a hypothetical (not yet on the market) health insurance package (18). This approach has been used because it has been proven to be an effective way of assessing willingness to pay with minimal bias for goods that are not presently available on the market. The CVM creates a hypothetical marketplace in which no actual transactions are made; hence, this approach has been successfully used for commodities that are not exchanged in regular markets. This approach makes the assumption that people have had no previous experience buying the health service that is going to be put on the market and instead asks people their willingness to pay on the basis of their expectations and the benefits attached to the service (19). In the absence of markets for public goods, this method presents consumers with hypothetical markets in which they have the (hypothetical) opportunity to buy the good in question. Since the elicited WTP values are contingent upon the particular hypothetical market described to the respondent, this approach is called the contingent valuation method (20).

### Study setting

The study was conducted in the Iganga and Mayuge districts of eastern Uganda. Iganga and Mayuge districts were selected purposively because they are the most poverty-hit districts in the country and 80% of the population lives in rural areas. They also have 80–85% of the population belonging to the informal sector (7). Iganga District is bordered by Kaliro District to the north, Bugweri District to the east, Mayuge District to the south, Jinja District to the southwest, and Luuka District to the west. With a total population of 504,197 people, 9.9% of the households are 5 km or more from the nearest health facility, and 19.9% are near a public health facility (UBOS, 2016). It is composed of 3 counties, 8 sub counties, 66 parishes, and 395 villages. The major economic activity is farming. Mayuge district is located in south-eastern Uganda with a population of 473,2394 and a growth rate of 3.5%, and it has thirteen subcounties. The major economic activities are farming and fishing. Mayuge district is bordered by Lake Victoria to the south, as well as Jinja and Bugiri districts. It has a total of 33 landing sites where fishing activities take place.

### Study Population

The study targeted the four major categories of informal sector workers: 1) farmers; 2) fishermen; 3) commercial cyclists; and 4) the business community (traders and market vendors) in the study area. The informal sector was chosen because they make up the largest (80%) portion of the population, without which the sustainability of the scheme will be questionable.

### Sample Size

The sample size was calculated using the Kish Leslie formula (21), considering a 95% confidence interval, a 50% expected level of WTP, a 5% level of precision, a design effect of 2, and a 10% non-response rate. This gave a total sample size of 853 informal sector workers. Four focus group discussions were conducted with women (four FGDs), and three FGDs were conducted with men (in the informal sector). This number depends on the level of information saturation. This was determined by doing preliminary analysis while still in the field.

### Sampling Procedure

Stratified random sampling was used in this study. For Iganga district, the informal sector workers were divided into three major categories (strata): farmers, commercial cyclists, traders/market vendors. In Mayuge district, the informal sector workers were divided into four categories: fishermen, farmers, commercial cyclists, and market vendors. A random sample from each stratum was taken in a number proportional to the stratum’s size when compared to the population. In Iganga district, two subcounties were randomly selected, and five villages from each subcounty were also randomly selected. A proportionate sample of 286 households from the 10 villages was selected, and the household heads were systematically selected for interviews. Seven *bodaboda* stages located along main roads and junctions were randomly selected, and 10 cyclists available at the time of the study were randomly selected to participate in the interviews. A total of 84 traders were selected systematically from the two main markets for interviews. In Mayuge district, a total of 10/33 landing sites were randomly selected, and 248 fishermen were selected from the 10 landing sites proportionate to size. Sixty-two traders were randomly selected from three main markets systematically, and 62 bodaboda cyclists were randomly selected from six stages located along main roads in Mayuge town (Table 1). Men and women from each of the categories were purposively selected for the focus group discussions.

**Table 1:** Sampling procedure per category of informal sector.

| **Iganga district (Iganga district planning office, 2018)** |  |  |  |  |
| --- | --- | --- | --- | --- |
| **Category** | **Farmers/peasants** | **Commercial cyclists** | **Traders/vendors** | **Fisherfolk** |
| Proportion in the district | 0.65 | 0.16 | 0.19 |  |
| Sample size | $(0.65 \times 440) = 286$ | $(0.16 \times 440) = 70$ | $(0.19 \times 440) = 84$ | - |
| Selection procedure | 2 sub counties (SC) and 5 villages from each SC were randomly selected. Households proportionate to size selected. | 7 *Boda boda* stages in the town were randomly selected, cyclists on stage randomly selected to be interviewed | Traders were systematically selected for the interviews two main markets | - |
| Mayuge district |  |  |  |  |
| **Category** | **Farmers/peasants** | **Commercial cyclists** | **Traders/vendors** | **Fisherfolk** |
| Proportion in the district | 0.1 | 0.15 | 0.15 | 0.6 |
| Sample size | $(0.1 \times 413) = 41$ | $(0.15 \times 413) = 62$ | $(0.15 \times 413) = 62$ | $(0.6 \times 413) = 248$ |
| Selection procedure | One sub county and two villages were randomly selected, the number of households were determined proportionate to size. | 6 *Boda boda* stages in town were randomly selected, later cyclists on stage at that time were randomly selected to be interviewed | Traders were systematically selected from 3 main markets | 10/33 landing sites were randomly selected. Fishermen at each site were randomly selected, proportionate to size |
| **Total samples** | **327** | **132** | **146** | **248** |

### Data collection methods and tools

The willingness to pay was assessed using a contingent valuation method using the “bidding game” technique. The bidding game was used to determine the maximum premium that a respondent is willing to pay for the proposed NHI plan. A bidding game technique was chosen because of its ease of implementation and its minimal biases. In addition, many studies have found the bidding game to be very reliable (22, 23). The respondent was asked if he or she was willing to pay the starting bid. If the respondent agreed, the interviewer would raise the bid by 10% of the first bid and again ask if the respondent was willing to pay the new bid. The interviewer continued to present the different bids until the respondent expressed an unwillingness to pay. On the other hand, if the respondent expressed unwillingness to pay the starting bid, the interviewer lowered the bid by 10% and repeated the query. This was continued until a bid was reached where the respondent was willing to pay. The starting bid was 20,000 UGX (5.4 USD), since this was the average premium for beneficiaries in the existing community-based health insurance schemes in Luweero, Western Uganda, and Jinja. The proportion of those willing to pay, the average, and the median amount of money people were willing to pay were computed. The perceived ability to pay was measured by asking respondents whether they were able to pay premiums for health insurance per year and how much they could afford without foregoing other basic needs.

A semi-structured questionnaire was used to collect quantitative data such as sociodemographics, WTP, perceived ability to pay, and associated factors from informal sector workers. The data was collected in April and May 2019. The data collection tools were translated to the local language and then back translated to English to check whether the translated questions still held informational validity. The principal investigator recruited and trained research assistants so that they were familiar with the statement of the problem, objectives of the study, sampling procedure, data collection tools, and plan for data collection. Research assistants were oriented to the bidding game technique for assessing WTP. They were also taught the meaning of health insurance and how it intends to be operationalized in the country. Research assistants were also trained on the basic interview techniques, such as asking questions in a neutral manner, not showing by words or actions what answers were being expected of the study participants, and how to record answers, especially from open-ended questions, without interpreting them. The data collection tools were pre-tested in the neighboring district, Bugweri district. This was done to test the validity and ease of application of the tools. Filled electronic forms were checked at the point of data collection for completeness, and those found incomplete were completed before the respondent was discharged. The data were cleaned and edited by the research assistants before they submitted the complete forms to the server.

Focus group discussion (FGD) guides were also used to collect data from informal sector workers to understand the determinants of willingness to pay and their perceived ability to pay for the NHI. The FGDs were audio-taped. The FGD was composed of 6–8 participants. The discussion had a moderator and a note-taker, and the discussions lasted for an average of one hour. These were conducted in the local language, and these were audio recorded.

### Data management and analysis

The quantitative data were downloaded from the server and imported into STATA 14 for cleaning and analysis. Descriptive statistics, including mean, median, standard deviations, frequencies, and percentages, were obtained. The information was presented in frequency distribution tables. Principal component analysis was done to generate wealth quintiles using the demographic health survey categorization. The wealth quintiles were generated based on nine household items, which included possession of a car, motorcycle, bicycle, radio, television, mobile phone, piece of land, owning a house, and having animals. Inferential statistics were obtained using logistic regression. This was done in two stages: 1) bivariate analysis was done to determine the potential variables associated with willingness to pay for health insurance. Crude odds ratios at a 95% confidence interval were obtained to measure the potential association. 2) A multivariate analysis was done to determine the actual factors associated with WTP. All variables that were statistically significant at bivariate analysis and the important factors known in the literature to be statistically significant, such as socio-economic status, occupation, and potential confounders like residence, were included in the model. Variables were tested for multicollinearity before being included in the model. The model was built step-by-step while testing it using lfit until the best model was obtained. Variables that remained statistically significant at 95% confidence were considered the factors associated with WTP for health insurance. Adjusted odds ratios were obtained as measures of association.

Qualitative data was analyzed manually using a thematic-inductive approach. The recordings were transcribed verbatim and translated from the local language (Lusoga) to English. Transcripts were read by more than two independent analysts who identified codes, sub-themes, and then organized them into themes. The information was triangulated with the quantitative data findings to gain a deeper understanding.

### Ethics statement

The study was approved by the Makerere University, School of Public Health Higher Degrees, Research and Ethics Committee (HDREC). At the district level, permission to conduct the study was obtained from the local leaders and the District Health Office. Permission was also sought from subcounty chiefs and local council one chairpersons. The informed written consent of each individual participant was obtained at the start of the study. Confidentiality and anonymity were maintained by the use of codes.

## Results

### Background Characteristics of the Respondents

Of the 853 respondents who participated in the study, 327 (38.3%) were peasant farmers, 248929.1%) were fishermen, 146 (17.1%) were business people, and 132 (15.5%) were commercial cyclists. The mean age of the respondents was 37.0 years (SD ± 11.3). The majority 667/853 (78.2%) of the respondents were males, 783/853 (91.8%) of the household heads were males, and 618/853 (72.4%) resided in rural areas. The average household size was 5 persons. The median monthly income was UGX 150,000 (USD 40.5). Only 158/853 (18.5%) were in the wealthiest quintile, while 349/853 (41.0%) of the households were in the second and poorest wealth quintiles (Table 2).

**Table 2:**
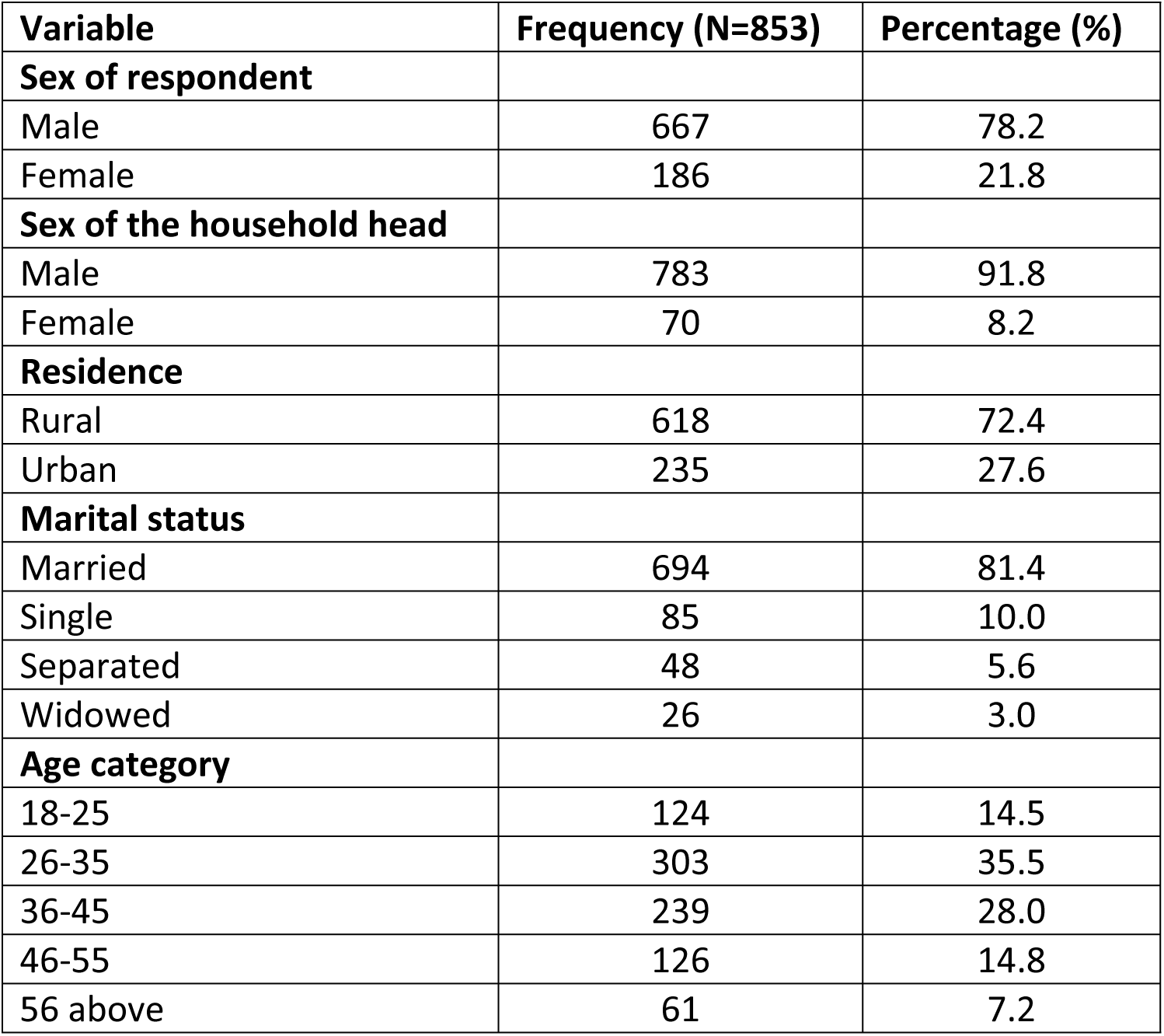
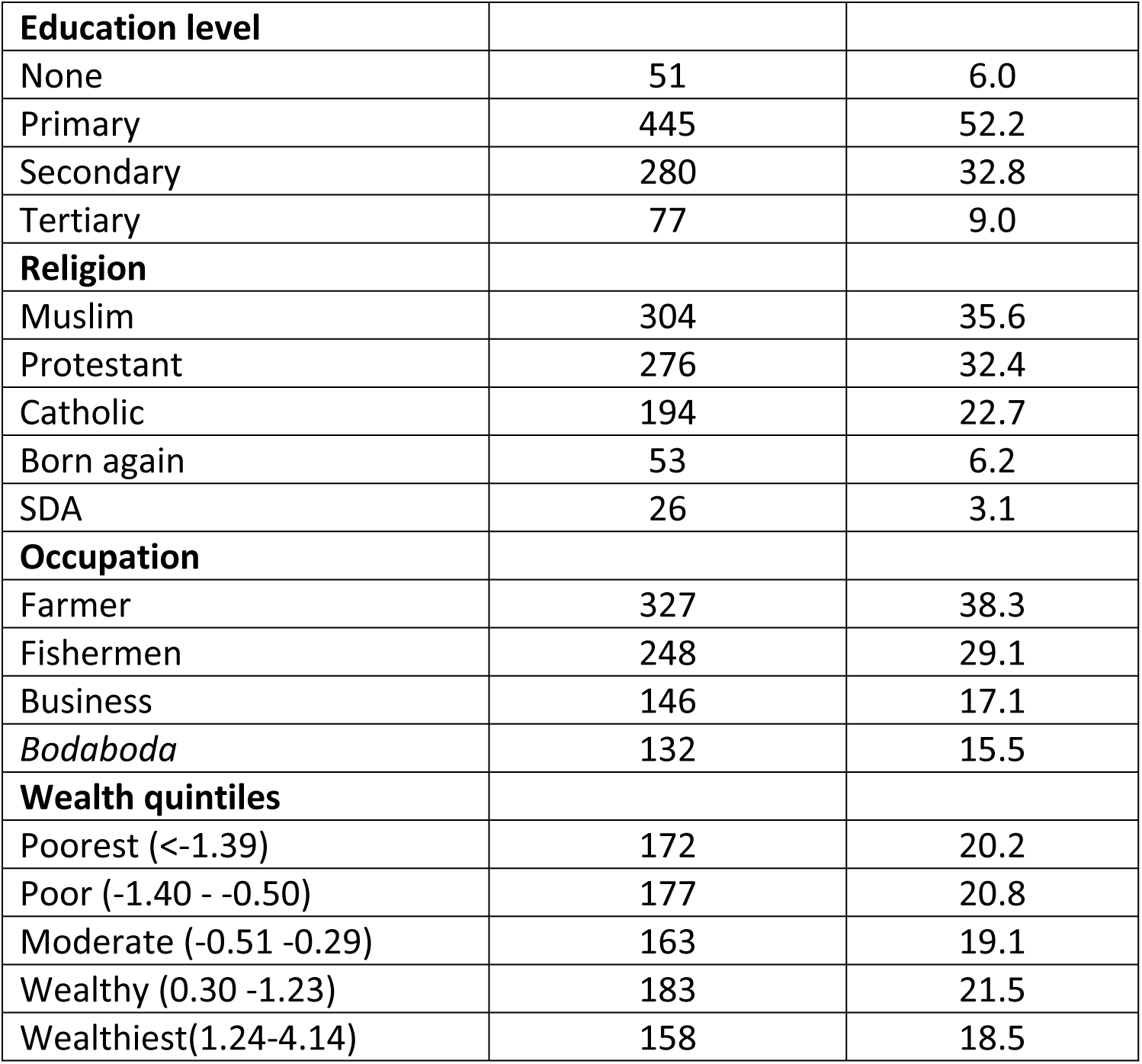
Background Characteristics of the Respondents.

### Willingness to Pay for the Proposed NHI

The majority 695/853 (81.5%) of the respondents were willing to pay for the proposed NHI. Of those willing to pay, the median willingness to pay was UGX 25,000 (USD 6.8) while the mean WTP was UGX 28,950 (USD 7.8) per person, inclusive of 4 dependents per year. Only 3.8% protested contributing to the scheme, and 126/853 (14.7%) gave WTP zero because they believed they were too poor to pay for national health insurance (Table 3).

**Table 3:** Willingness to Pay for the Proposed NHI annually.

| WTP | Frequency (%) | Mean | Median |
| --- | --- | --- | --- |
| Protests (WTP=0) | 32 (3.8) | 0 | 0 |
| WTP=0 | 126 (14.7) | 0 | 0 |
| WTP>0 | 695(81.5) | **28,950** | **25,000** |
| Total | 853 (100.0) | 23,600 | 20,000 |
| **Copayment scenario** |  |  |  |
| Amount willing to copay | 309/695(44.5) | **2,650** | **2,000** |
| WTP with co payment | 309/695(44.5) | 23,000 | 20,000 |

**Notes;** Protests-those protested attaching valuing to HI (WTP=0 but it has no economic sense).
WTP=0 means willingness to pay is actually zero. WTP>0 that it is positive -WTP is above zero.

Qualitatively, most of the FGD participants were also willing to make contributions for health insurance. However, others resented making contributions, as noted below:

> “Sincerely, the government is meant to provide all the medical services. So, there is no way that you can lie to us and say that the government can’t provide those services…” **(FGD male-fishermen-Mayuge)**

A number of FGD participants were not willing to pay because of poverty, while others criticized the government’s misuse of taxes collected from the tax payer on salaries for officials rather than on health services, as noted in the quotes below.

> “The fact is that poverty is the biggest hindrance. Because at times you can fall sick, yet you don’t have money, and you resort to borrowing from colleagues who may not even be able to help you out because they also don’t have. So generally, I disagree with paying for health insurance.” **(FGD-Business Women, Mayuge District)**

> *“I am not willing to pay because we are always charged a lot of tax to register our businesses. That is all tax. Where does that tax go? Let them reduce the salary of Members of Parliament so that part of it is utilized to buy medicine. Members of Parliament have no importance in our area.”* (**FFGD-Male-fishermen-Mayuge district)**

## Willingness to Pay with Copayments

A copayment is a cost-sharing arrangement where one covers a portion of the costs out of pocket each time one visits the health facility. Less than half, 309/695 (44.5%) of the respondents, were willing to co-pay. The mean co-pay amount was UGX 2,650 (USD 0.7), with a median of UGX 2000 (USD 0.5), each time someone accessed services. The average maximum amount of money that respondents were willing to pay with the copayment was UGX 23,000 (USD.6.2) and the median was UGX 20,000 (USD.5.4) (Table 3).

> “I also do not support the idea that we pay money each time we visit the health facility, especially when we are paying every year. This will make people unable to access the services, and hence the problem of access to services will not have been solved. People are already poor, and that will mean people will continue to sell their property to go to health facilities.” (FGD Male Farmer)

### Preferred Mode of Payment

Most (54.1%) respondents preferred to pay annually, while 21.0% preferred to pay per harvesting season. Only 13.1% wanted to pay according to their monthly income, with the majority (63.1%) preferring to pay 1 percent of their monthly income, followed by those who were willing to pay 5%.

### Perceived Ability to Pay for the Health Insurance Scheme

More than half 633/853 (74.2%) of the respondents believed that they were able to pay for health insurance, with a mean of UGX 23,000 (USD.6.2) and median of UGX 20,000 (USD.5.4) per person per year.

Regarding the perceived ability to pay, most of the FGD participants noted that most of the people are poor and most likely may not be able to make periodic subscriptions, especially if the money is raised more than what people earn. This was attested to, as indicated below:

> “Sincerely speaking, some people cannot afford to pay money; for instance, you may find a very old person who is no longer earning anything, yet they also need health care.” **(FGD-Boda-boda-Iganga district)**

Another FGD participant, who was a business lady, noted that

> “I support the idea. But not very much because of the issue of affordability. Not everyone can afford to pay, and note that we earn differently. Because some of us play the role of mother and father in the family, there are a lot of requirements to provide.” **(FGD Business Women, Iganga)**

Furthermore, most of the participants noted that setting premiums should be based on people’s income and ability to pay, as noted below:

> “Since everyone earns differently, the charges should depend on how much someone earns. I could be in a position to afford UGX 50,000, but yet my colleague cannot at all since people in the village are used to getting free medicine. When some people are asked to pay just 500shs for drugs, they will consider it unworthy because they know that drugs/medication are freely given. So, if possible, we charge according to people’s income.” **(FGD: business women, Iganga district)**.

### Determinants for level of Willingness to Pay for Health Insurance Scheme

At bi-variable analysis, wealth quintiles, having savings group membership, having a family member with chronic illness, use of traditional medicine, and hearing about health insurance were statistically significant for willingness to pay for health insurance. The odds of being willing to pay when in the fourth wealth quintile were 2.88, compared to the odds of being willing to pay among those in the poorest category (COR: 2.88, 95% CI: 1.60–5.18, P<0.001). Not being in the saving group reduced the chances of being willing to pay for health insurance by 42% compared to those who had saving group memberships (COR: 0.58, 95% CI: 0.33-0.69, P<0.001) (Table 4).

**Table 4:**
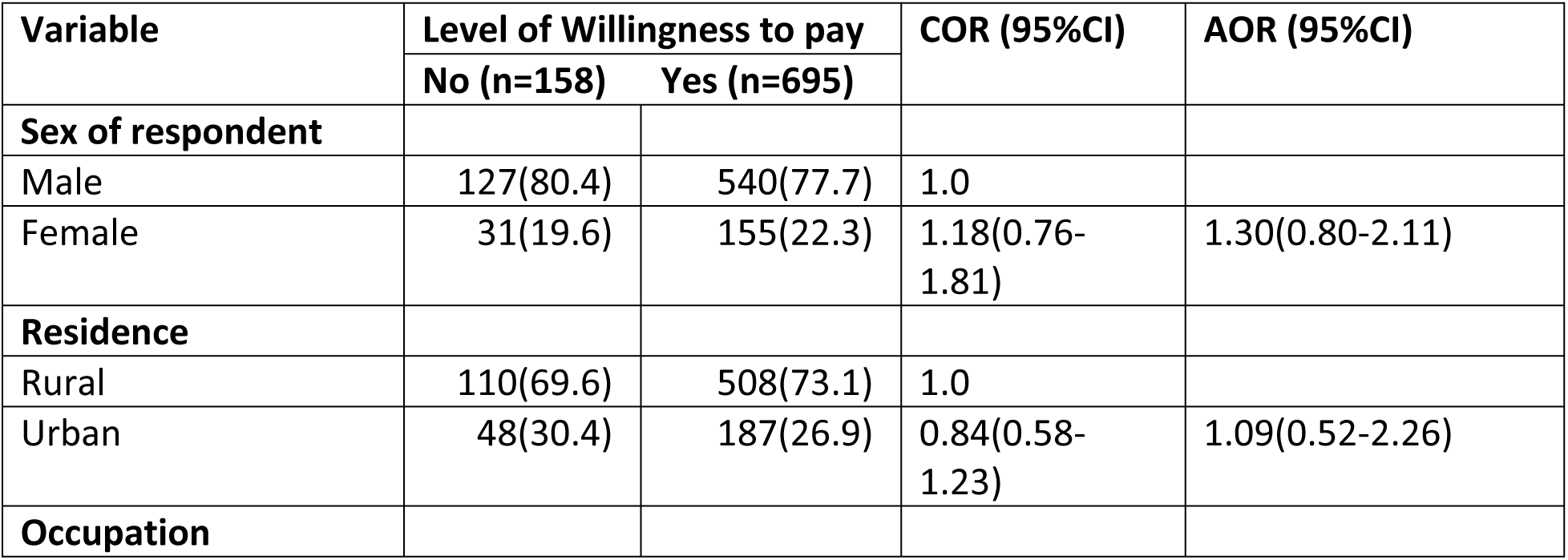
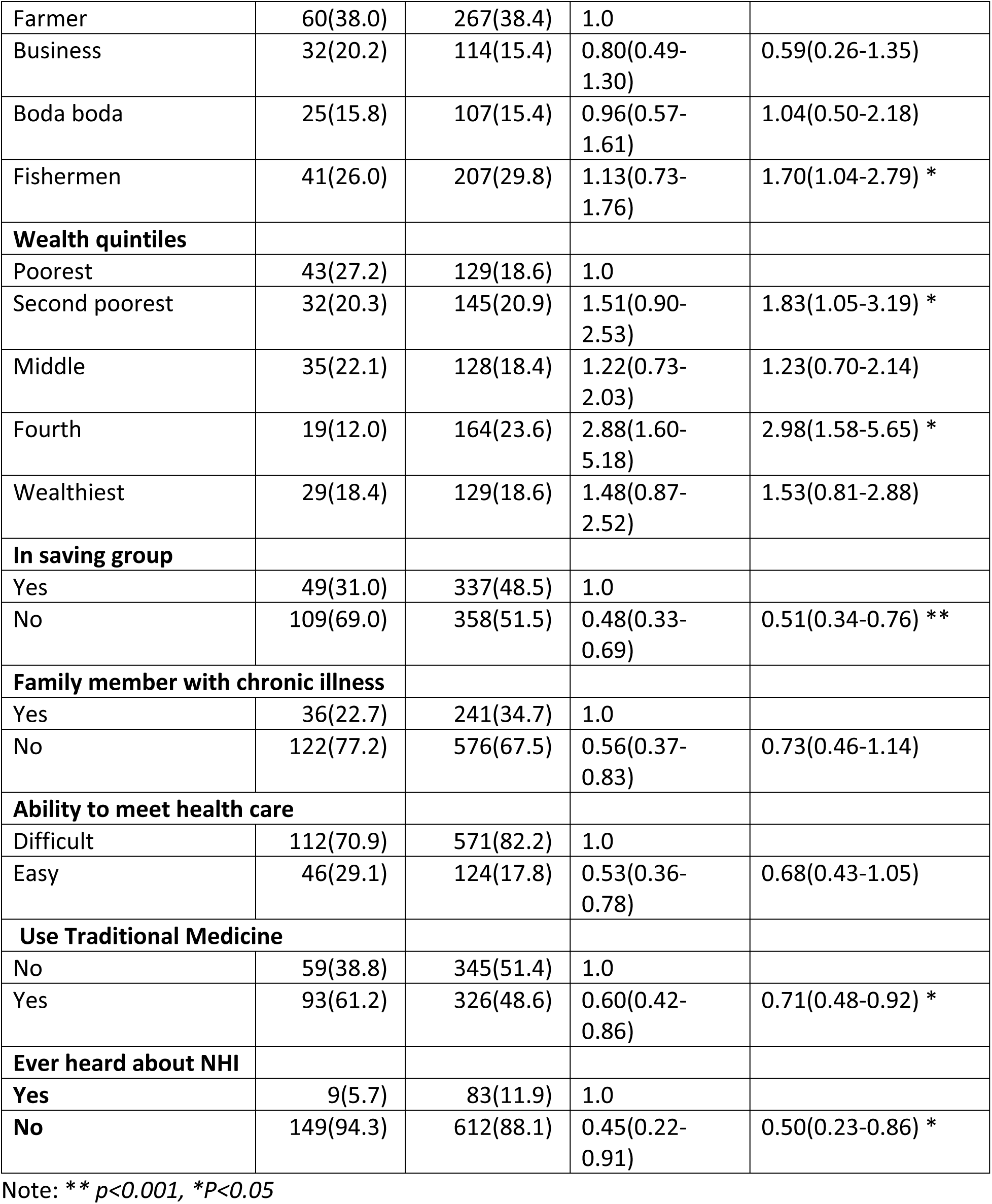
Multi variable analysis of factors associated with WTP for NHI scheme.

After adjusting for any confounding factors, occupation, wealth, saving group membership, use of traditional medicine, and having heard about health insurance were significantly associated with the level of willingness to pay for the proposed health insurance scheme.

The odds of fishermen’s willingness to pay for health insurance were 1.70 compared to those of farmers after adjusting for other factors (AOR: 1.70, 95% CI: 1.04–2.79, P = 0.035). Respondents in the fourth wealth quintile were 2.98 times more likely to pay for health insurance compared to those in the poorest wealth quintile (AOR: 2.98, 95% CI: 1.56–5.65). Respondents who were not in any saving group were 0.51 times less likely to pay for the proposed health insurance compared to those with saving group membership (AOR: 0.51, 95% CI: 0.34-0.76, P<0.001), and individuals who had never heard about HI were 50% less willing to pay for the proposed National Health Insurance compared to those who had ever heard about health insurance (AOR: 0.50, 95% CI: 0.23-0.86, P = 0.032) (Table 4).

## Discussions

Firstly, this study is unique in its assessment of the level of WTP, perceived ability to pay, and determinants of WTP for the proposed national health insurance scheme among the informal sector workers in Uganda, who constitute 80% of the Ugandan population. These findings are pertinent if the proposed NHI is to achieve adequate coverage. Secondly, the study applied a contingent valuation approach using a bidding game technique to hypothetically elicit the informal sector’s willingness to pay. The highlight of this study is the high level of willingness to pay for NHI among informal sector workers, as well as their preferred modes of engagement. These findings provide valuable insights for fine-tuning the proposed NHI.

The level of WTP for the proposed NHI was high, with a median willingness to pay of UGX 25,000 (USD 6.8) and a mean WTP of UGX 28,950 (USD 7.8) per person, inclusive of 4 dependents per year. This level of willingness to pay was higher than the reported WTP in Nepal (71%) and in South Sudan (68%) among the informal sector (13, 24). However, the maximum amount of money people were willing to pay was lower than the 5.5 USD per person per month in Iran, 6.6 USD per person per month in Namibia, and 3 USD in Kenya per month (18, 24, 25). The differences in WTP across the different countries could be due to the differences in the socio-economic status of the different populations in the different countries. The high level of willingness to contribute to health insurance highlights high chances of the scheme being successful. However, the suggested premium being higher than the stated WTP may hinder many from participating in the scheme. But this also brings up the policy question of whether the WTP can sustain the scheme. Thus, there is a need for careful revision of the premiums before Parliament passes the NHI bill.

The ability to pay is another important consideration when planning to implement health insurance, especially among the informal sector, whose incomes are unpredictable. A high number of participants believed that they could afford to make periodic contributions to the health insurance scheme in this study. This finding further highlights the potential for the NHI to succeed if the informal sector is included. However, the ability to pay needs to be considered when setting premiums, given the fact that less than 20% of the studied population were in the wealthiest category.

Although more than half of the respondents earned more than 100,000 UGX (27 USD) a month, there are substantial sections of the informal sector that will struggle to pay for health insurance, and therefore, these will require significant government subsidies to be included. This further highlights the need to consider the idea of making contributions as a proportion of people’s earnings and wealth status because it would make people pay according to their ability and is more equitable than just having a flat figure for all people. Moreover, by virtue of the unpredictable nature of their income, informal sector workers expressed the desire to have premiums synchronized to earning cycles such as harvest seasons. This implies that the NHI will have to explore flexible premium schedules for informal sector workers in order to enhance their uptake of the scheme. In Ethiopia, making premium systems more flexible eased payments (26).

In this study, having a savings group membership increased the likelihood of willingness to pay for the proposed national health insurance scheme. In Asian countries, saving groups were reported to be key in initiating community-based health insurance schemes (27). Savings groups provide members with a secure place to save money, generate a pool fund, and have the opportunity to borrow in small amounts. They also provide affordable basic insurance services and enable the community to meet the premiums(27). Local savings groups have become one of the most famous financial support systems in rural Uganda over the last decade (28). Savings and burial groups present an opportunity to leverage in enhancing access to health care in rural communities. This implies that as countries plan to initiate NHI, saving groups can become a great resource to tap into to ensure community participation and payment of the premiums for the scheme.

Participants belonging to the highest wealth category were more likely to pay for the proposed national health insurance scheme. This is in agreement with studies conducted in Nigeria, Ethiopia, and Ghana, which have reported that socio-economic factors such as poverty, income, or wealth status have a bearing on the ease or difficulty of enrolling in a given scheme, with high premiums deterring enrollment in schemes (29–31). Similar studies conducted in Sierra Leone and Iran showed that the WTP for the HI scheme depended on the monthly income of the respondent (25, 32). In Nepal, belonging to a lower wealth quartile was associated with a lower willingness to pay for National Health Insurance Scheme (33). This therefore underscores the need for income generating projects to be implemented within the communities to aid them make contributions for health insurance.

The use of traditional medicine was negatively associated with participation in the health insurance scheme. This is because members who use traditional medicine see no need to utilize modern health care services, making them less likely to pay for health insurance. Studies have shown that the presence and use of alternative medicine and other forms of healthcare, such as herbalists, negatively affects participation in a health insurance scheme since insurance requires regular use of modern medicine. Studies conducted in Cameroon, Burkina Faso, Nigeria, and India have reported that those who use modern medicine are more willing to pay for health insurance than those who use traditional medicine (34–36). In Uganda’s pluralistic health system, the ministry of health and other mandated regulatory bodies need to institute measures that not only regulate the alternative sector but ultimately attract people to use modern medicine as opposed to traditional medicine.

It is not surprising that those who had never heard about health insurance had 50% less WTP compared to those who had prior information about health insurance. Indeed, the level of knowledge on how health insurance operates was very low, and statistical tests of significance could not be used in this study. However, one systematic review and meta-analysis of factors influencing WTP for voluntary contributary health insurance schemes in LMICs has revealed that knowledge and understanding of the functioning of the scheme positively influence the willingness to pay for and participate in the scheme (37, 38). As the Ugandan government moves to implement health insurance, information, education, and communication will be paramount to the success of the proposed scheme.

## Study Limitations

As with most contingent valuation studies, the elicitation technique is always subject to bias, and the assignment of the first bid or amount is also biased. However, this was triangulated with an open-ended question and qualitative data. The low level of knowledge about health insurance among respondents was also a limitation for attaching value to the proposed scheme. This was solved by educating the respondent about health insurance before eliciting a willingness to pay.

## Conclusions

This study indicated a high level of WTP, and the majority believed they could afford to pay for the health insurance scheme. However, studies on the ability to pay need to be conducted, as this study is based on the perceived ability to pay for health insurance schemes. The high level of WTP indicates that the informal sector is willing to make a contribution to their health, making it feasible for the government to propose a flexible and affordable scheme within which the informal sector can bear some costs towards their health care. To support the ability to pay, there is a need to build local solidarity groups and strengthen local initiatives to aid poor members.

## Declarations

### Consent for publication

Not Applicable

### Availability of data and materials

The datasets analyzed for this study are available from the corresponding author on reasonable request.

### Authors’ contributions

NN, SNK, AK conceived the study, NN collected, analyzed and wrote the first draft of the manuscript. SNK and AK provided reviews on all drafts of manuscript. All the authors read and approved the final manuscript.

## Acknowledgements

We wish to thank the District Health Officers of Mayuge and Iganga districts for their cooperation, mobilization and ensuring that data collection went on smoothly. We also extend our sincere appreciation to the research assistants and all respondents in Iganga and Mayuge for sparing their time to contribute to this piece of work.

